# Epidemiological, socio-demographic and clinical features of the early phase of the COVID-19 epidemic in Ecuador

**DOI:** 10.1101/2020.05.08.20095943

**Authors:** Esteban Ortiz-Prado, Katherine Simbaña-Rivera, Ana Maria Diaz, Alejandra Barreto, Carla Moyano, Vannesa Arcos, Eduardo Vásconez-González, Clara Paz, Fernanda Simbaña-Guaycha, Martin Molestina-Luzuriaga, Raúl Fernández-Naranjo, Javier Feijoo, Aquiles R. Henriquez, Lila Adana, Andrés López Cortés, Isabel Fletcher, Rachel Lowe, Lenin Gómez-Barreno

**Author notes:** **Corresponding author:** Esteban Ortiz-Prado One Health Research Group, Universidad de las Américas, Quito, Ecuador Calle de los Colimes y Avenida De los Granados, Quito 170137, Ecuador.

## Abstract

**Background:** The SARS-CoV-2 virus has spread rapidly around the globe. Nevertheless, there is limited information describing the characteristics and outcomes of COVID-19 patients in Latin America.

**Methods:** We conducted a cross-sectional analysis of 9,468 confirmed COVID-19 cases reported in Ecuador. We calculated overall incidence, mortality, case fatality rates, disability adjusted life years, attack and crude mortality rates, as well as relative risk and relative odds of death, adjusted for age, sex and presence of comorbidities.

**Results:** A total of 9,468 positive COVID-19 cases and 474 deaths were included in the analysis. Men accounted for 55.4% (n = 5, 247) of cases and women for 44.6% (n = 4, 221). We found the presence of comorbidities, being male and older than 65 years were important determinants of mortality. Coastal regions were most affected by COVID-19, with higher mortality rates than the highlands. Fatigue was reported in 53.2% of the patients, followed by headache (43%), dry cough (41.7%), ageusia (37.1%) and anosmia (36.1%).

**Conclusion:** We present the first analysis of the burden of COVID-19 in Ecuador. Our findings show that men are at higher risk of dying from COVID-19 than women, and risk increases with age and the presence of comorbidities. We also found that blue-collar workers and the unemployed are at greater risk of dying. These early observations offer clinical insights for the medical community to help improve patient care and for public health officials to strengthen Ecuador’s response to the outbreak.

## Introduction

For the past few decades, the world has been exposed to a series of threats from viral outbreaks caused by emerging zoonotic diseases and in particular by a family of viruses known as coronaviruses^1^. The World Health Organization recognizes at least three types of coronavirus capable of generating epidemic outbreaks, including SARS-CoV, MERS-CoV and the recently discovered SARS-CoV-2 virus^2^. These viruses are responsible for causing severe acute respiratory syndrome (SARS), Middle East respiratory syndrome (MERS) and the most recently described coronavirus disease (COVID-19)^3^.

Since the first reports of a cluster of atypical pneumonia cases in Wuhan, China on December 2019, the SARS-CoV-2 virus and COVID-19 has quickly spread across the globe, infecting more than 3,585,936 people and causing more than 245,803 deaths worldwide ^1,4^ One of the main reasons the virus has spread so rapidly is due to its airborne nature of transmission from both symptomatic and asymptomatic people, making it difficult to test, trace and isolate new cases effectively ^2^

Understanding the transmission dynamics in different settings can provide important clues about the advance of the pandemic, especially in areas with unequal access to health services, high population density and a high burden of neglected tropical diseases ^5,6^.

In this study we present the findings of an interim analysis of the epidemiological situation in Ecuador, describing the clinical characteristics and epidemiological behavior of the first 9.468 laboratory confirmed COVID-19 cases officially registered in Ecuador.

## Methods

We conducted a country-wide population-based analysis of the epidemiology of the first 9.468 COVID-19 patients reported in Ecuador between 27 February and 18 April 2020 (Figure S1-Appendix 1). We obtained socio-demographic variables, such as age, sex, marital status and place of residence from the Ministry of Health (MoH) registries. Clinical data including date of onset of symptoms, date of diagnosis and date of death, as well as the presence of comorbidities, pregnancy and influenza vaccination history were also obtained. Epidemiological information including city and province of registration, elevation, occupation, travel history and institution of diagnosis were analyzed.

The attack rate, mortality rate, and case fatality rate were computed using the population at risk living in a canton or a province. CFR% were adjusted to mitigate the effect in crude CFR% estimation. We computed the time distribution from hospital admission to the time of using the methodology reported by Russel 2020^7^.

Relative risks (RR) and 95% confidence intervals were computed for all age groups using female cases as the reference level. We performed a relative risk (RR) analysis of the total number of expected cases by the population at risk in each group to obtain the likelihood of dying due to COVID-19. In order to control for the effects of sex, age and comorbidities, an adjusted logistic regression was performed using the final outcome (recovered or died) as a response variable.

The number of years of healthy life lost due to COVID-19 among the reported cases were calculated using Disability-Adjusted Life Years (DALY). DALYs are the sum of the Years of Life Lost (YLL) due to premature mortality in the population and the Years Lost due to Disability (YLD) caused by the consequences of the disease ^8,9^

## Ethical consideration

This secondary data analysis of anonymized, un-identifiable information received ethical exemption from the Universidad de las Americas Ethics Committee CEISH on March 10^th^, 2020 (Appendix 1).

## Results

There were 9,468 positive cases of COVID-19 and 474 officially reported deaths in Ecuador from 27 February – 18 April 2020 (54-day period). 99.3% of COVID-19 patients were Ecuadorians (n = 9,400), 0.30% (n = 44) were from other countries in Latin America and the other 0.40% (n = 24) were either from Europe, North America or Asia (Figure 1).

**Figure 1.**
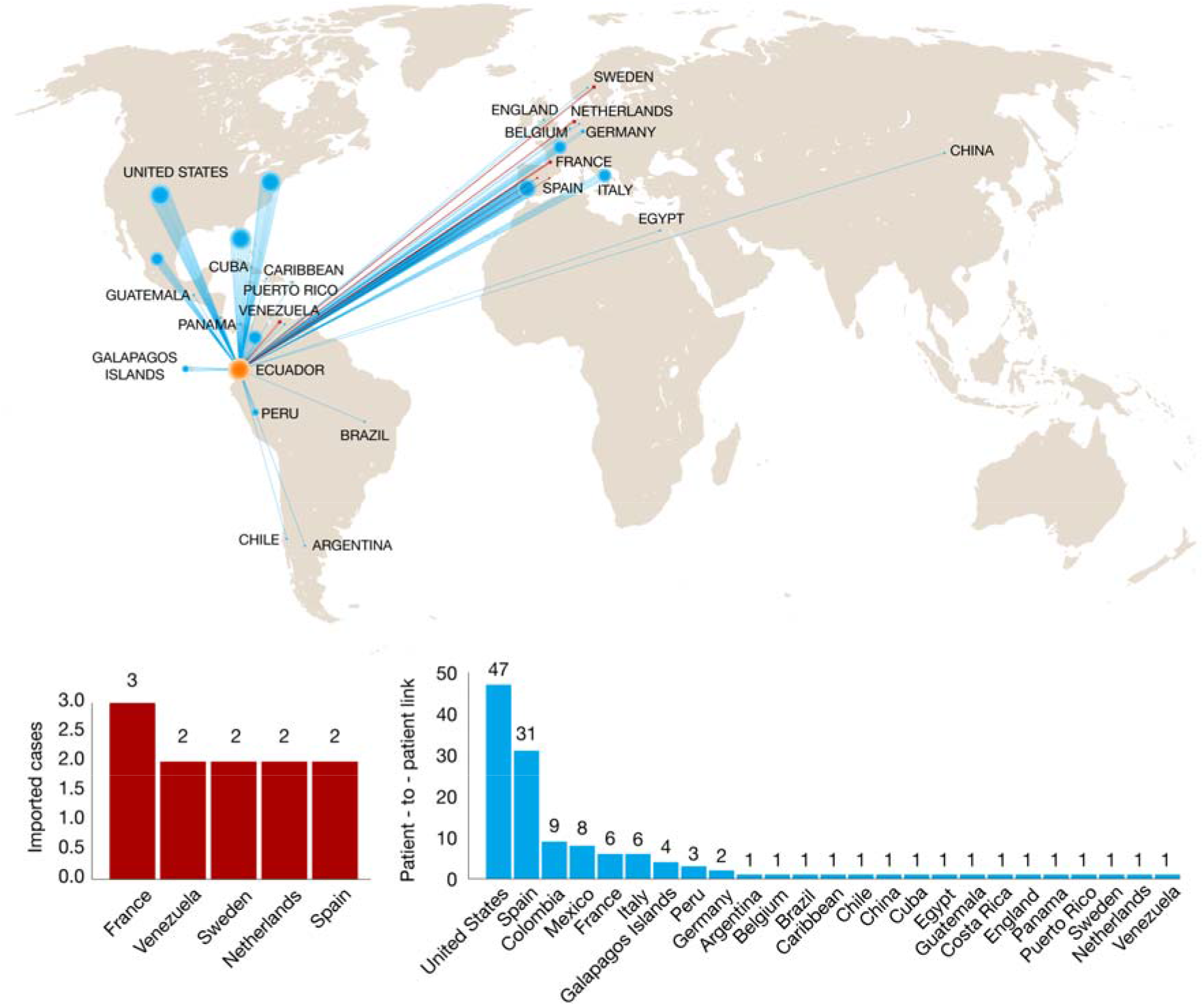
Origin of imported (n = 11) (red bars) and community-transmitted cases (n = 132) (blue bars) with direct travel history among COVID-19 confirmed patients in Ecuador.

Men accounted for 55.4% (n = 5,247) of all cases with an incidence rate of 60.5 per 100,000, while women accounted for 44.6% of cases (n = 4,221) and an incidence of 47.2 per 100,000. The median age of COVID-19 patients was 42 (IQR: 32-56) in men and 39 (IQR: 30-54) in women. The median age of patients who had died from COVID-19 was 62 (IQR: 51-70) in men and 65 (IQR: 56-74) in women (Figure 2, Supplementary Table 1). We also found that men were more likely to die from COVID-19 in almost every age group (Figure 1), although case fatality rate for patients aged between 85-94 was higher amongst women.

**Figure 2.**
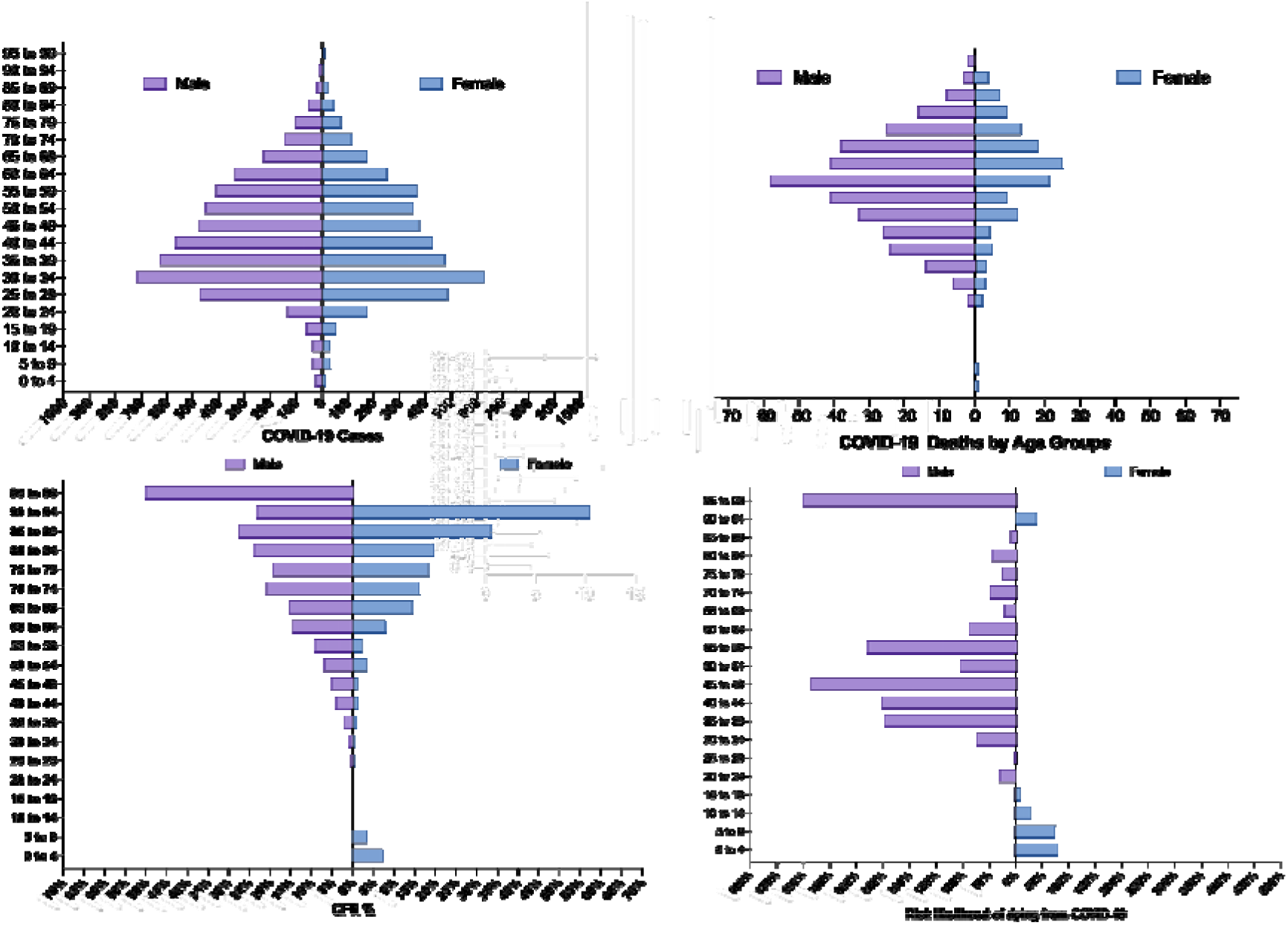
COVID-19 cases and deaths in Ecuador by age and sex. Top panel: Age pyramids for confirmed cases and officially reported deaths. Lower panel: Case fatality rate (%) and risk likelihood of dying from COVID-19, for females (blue) and males (purple).

The majority of COVID-19 cases occurred among Mestizos 78% (n = 7,367), followed by indigenous 0.79% (n = 75), Caucasians 0.84% (n = 40) and Afro-Ecuadorians/Black with ~0.1% (n = 16) (Table 1).

**Table 1.** Number deaths, patients recovered and case fatality of COVID-19 in Ecuador. Total number of deaths, patients recovered and percentage of total (%) and case fatality rate (%) for women and men in different ethnic groups, type of healthcare provision, presence of comorbidities and travel history.

|  | **Women** |  |  | **Men** |  |  |
| --- | --- | --- | --- | --- | --- | --- |
|  | Deaths<br>(n = 137) | Recovered<br>(n = 4,084) | CFR (%) | Deaths<br>(n = 337) | Recovered<br>(n = 4,910) | CFR (%) |
| **Ethnicity (% of population)** |  |  |  |  |  |  |
| Afro Ecuadorian<br>(7.2%) | 0 (0%) | 4 (0.1%) | 0% | 0 (0%) | 2 (0%) | 0% |
| Caucasian (6.1%) | 1 (0.7%) | 16 (0.4%) | 5.9% | 0 (0%) | 23 (0.5%) | 0% |
| Indigenous (7%) | 2 (1.5%) | 30 (0.7%) | 6.3% | 5 (1.5%) | 38 (0.8%) | 11.6% |
| Mestizo/a<br>(71.9%) | 128 (93.4%) | 3,197 (78.3%) | 3.8% | 314 (93.2%) | 3,728 (75.9%) | 7.8% |
| Montubio (7.4%) | 3 (2.2%) | 39 (1%) | 7.1% | 10 (3%) | 40 (0.8%) | 20% |
| Mulato | 0 | 3 (0.1%) | 0.0% | 0 (0%) | 2 (0%) | 0% |
| Black | 0 | 4 (0.1%) | 0.0% | 0 (0%) | 6 (0.1%) | 0% |
| Others (0.3%) | 1 (0.7%) | 8 (0.2%) | 11.1% | 0 (0%) | 6 (0.1%) | 0% |
| No data | 2 (1.5%) | 783 (19.2%) | 0.3% | 8 (2.4%) | 1,065 (21.7%) | 0.7% |
| **Healthcare provider** |  |  |  |  |  |  |
| Private | 6 (4%) | 995 (24.4%) | 0.6% | 12 (3.6%) | 1,424 (29%) | 0.8% |
| Public | 131 (96%) | 3,089 (75.6%) | 4.1% | 325 (96.4%) | 3,486 (71%) | 8.5% |
| **Presence of comorbidities** |  |  |  |  |  |  |
| Yes | 22 (16.1%) | 191 (4.7%) | 10.3% | 55 (16.3%) | 270 (5.5%) | 16.9% |
| No | 115 (83.9%) | 3,893 (95.3%) | 2.9% | 282 (83.7%) | 4,640 (94.5%) | 5.7% |
| **Travel history** |  |  |  |  |  |  |
| Yes | 2 (1.5%) | 66 (1.6%) | 2.9% | 9 (2.7%) | 64 (1.3%) | 12.3% |
| No | 55 (40.1%) | 1160 (28.4%) | 4.5% | 139 (41.2%) | 1496 (30.4%) | 8.5% |
| No data | 80 (58.4%) | 2,858 (70.0%) | 2.7% | 189 (56.1%) | 3350 (68.2%) | 5.3% |

These differences were compared with the national distribution for each ethnic group according to the 2010 national census ^10^. 19% of confirmed cases of COVID-19 in Ecuador (n = 1800) were amongst health professionals and from this group medical doctors (n = 876) were the most affected, representing 9.3% of all reported cases (Figure S3-Appendix 1).

Approximately 99% of patients with COVID-19 (n = 9,384) did not report any vaccination history in the last year and only 0.9% (n = 84) reported having been vaccinated. From those not vaccinated, 471 patients died whilst only three patients who had not been vaccinated died. Mortality risk among those not vaccinated was higher than patients who had been vaccinated, although the difference was not statistically significant (1.40 [95% CI: 0.46 to 4.28]). The relative risk of mortality in pregnant women was not found to be statistically significant (RR: 2.05 [0.785 to 5.36] p value: 0.14), although 6.7% of pregnant women in the study (n = 60) were reported to have died due to COVID-19.

Using an adjusted logistic regression model we found that the presence of comorbidities among COVID-19 patients, being male and older than 65 years old increased the mortality risk by almost 130% (OR: 2.27 [1.72-3.00, p value <0.001]), 100% (OR: 2.03[1.65-2.50, p value < 0.001]) and 470% (OR: 5.74 [4.7-7.0, p value <0.001]), respectively (Table 2).

**Table 2.** Results of adjusted logistic regression model for the effects of the presence of comorbidities, sex and age on mortality in COVID-19 patients. Parameter estimates, odds ratios, p-values and 95% upper and lower confidence intervals are shown

| Covariate | Parameter estimate | Odds ratio | 95% LCI | 95% UCI | p-value |
| --- | --- | --- | --- | --- | --- |
| **Presence of Comorbidities** |  |  |  |  |  |
| No (Reference) |  |  |  |  |  |
| Yes | 0.82 | 2.27 | 1.72 | 3.00 | <0.001 |
| **Sex** |  |  |  |  |  |
| Women (Reference) |  |  |  |  |  |
| Men | 0.71 | 2.03 | 1.65 | 2.50 | <0.001 |
| **Age** |  |  |  |  |  |
| <65 years |  |  |  |  |  |
| >65 years | 1.74 | 5.744 | 4.71 | 7.00 | <0.001 |
| Dependent variable: Death/Alive final outcome |  |  |  |  |  |

LCI: lower confidence interval; UCI: upper confidence interval

The most common symptom reported among COVID-19 patients in Ecuador was fatigue or general tiredness (53.2%), followed by headaches (43%), and dry cough (41.7%). 37.1% of the patients reported loss of taste (ageusia), 36.1% reported loss of smell (anosmia) and 35 % reported muscle and joint pain (Figure S2-Appendix 1). The median time elapsed between the onset of symptoms in COVID-19 patients and receiving medical attention was four days (IQR: 1-8 days). The average time between onset of symptoms in patients and case notification was nine days (IQR: 5-14 days) (Figure 3 and Appendix 1). When observed retrospectively using the date of the onset of symptoms, 29 undocumented patients were already sick but only 6 were diagnosed, trend that continued to be high until March 24^th^, the day when the lock-down was implemented in Ecuador (Figure 3).

**Figure 1.**
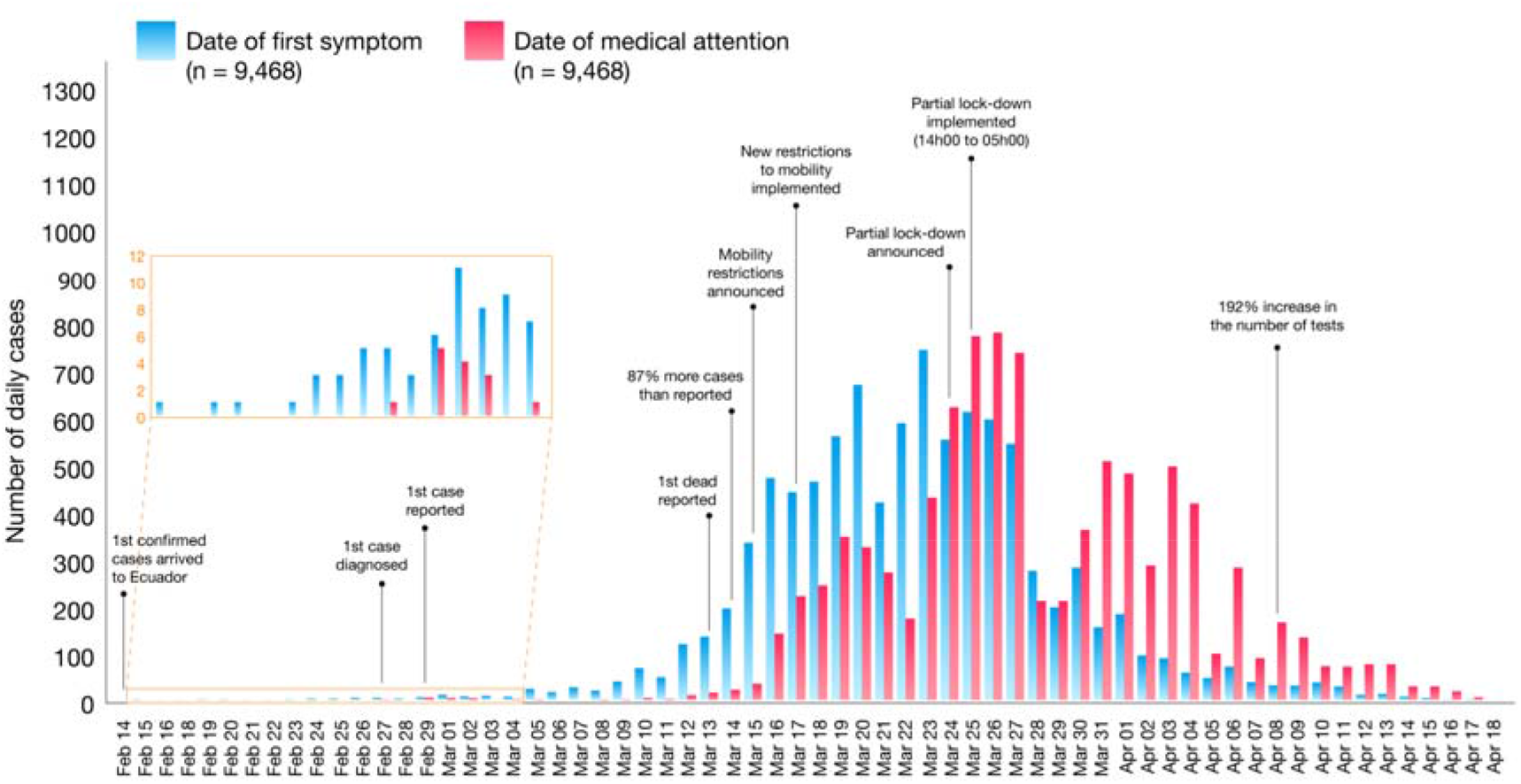
Epidemiological curves of COVID-19 cases by date of onset of symptoms (blue) and date of diagnosis (red), Ecuador from 14 February to 18 April 2020. The first case in Ecuador was diagnosed on February 27^th^ but notified on February 29^th^.

The overall attack rate of COVID-19 in Ecuador was 51.1 per 100,000 people. Sex-specific attack rates of COVID-19 were 60.5 per 100,000 for men and 47.2 per 100,000 for women. The lowest attack rate was amongst children from 0-4 years old (2.86 per 100,000), while the highest attack rate was found in patients aged between 55 and 59 years (111 per 100,000). Overall mortality rate for COVID-19 in Ecuador was 2.7 per 100,000 people, with patients aged 90-95 years being the most affected group with a crude age specific mortality rate of 20 per 100,000 people. There were 474 total deaths in Ecuador due to COVID-19, confirmed using RT-PCR. Between 1 March and 18 April 2020, 2,092 deaths related to acute respiratory distress syndrome were recorded as probable or suspected due to COVID-19 (Figure S5-Appendix 1).

We found that coastal regions had higher attack rates than the highlands (p-value: 0.011) and living above 2,500 m was associated with a lower risk of mortality (RR: 0.63 [CI 95% 0.50 - 0.79]), compared to populations living at lower altitudes (Figure S4, Figure S6, Figure S7-Appendix 1).

In terms of years of life lost prematurely (YLL), COVID-19 predominantly caused mortality among older adults, especially men. From the start of the outbreak at least 3,207 years were lost prematurely among women and more than 8,847 among men (Table 3). COVID-19 also caused a loss of 12,112 healthy life years, with an average of 1.27 DALY per case. From the estimated burden of COVID-19, 99.5% is attributable to years of life lost due to premature mortality, with an average of 25.4 YLL per death. The population in Ecuador between 20 to 64 years old contributed to 74.4% of disease burden, followed by the elderly with 24.2% of the burden (Appendix 1).

## Discussion

Ecuador has been the worst hit country in the Latin American region from the COVID-19 pandemic ^11^. Since the first case confirmed in Ecuador on 27 Feb 2020, at least 9,468 positive COVID-19 cases and 474 deaths were officially registered over a 54-day period.

The images of corpses in the streets and the difficulty of burying the dead occupied the main pages of all the newspapers around the world ^11,12^ Overcrowded hospitals and laboratories were overwhelmed by an excessive number of cases, causing a confirmed toll of 474 COVID-19 related deaths and 809 registered suspicious deaths up to April 18^th^ 2020 ^13^. On April 18, the National Emergency Operations Committee reported 1,061 patients discharged from hospital, 369 hospitalized patients, and 7,564 COVID-19 positives cases, who were stable in home isolation ^14^. Data from the region indicates that Ecuador, along with Panama, are the two most affected countries in Latin America. Ecuador ranks 23^rd^ worldwide in the numbers of deaths per million inhabitants ^15,16^.

At the beginning of the pandemic, Ecuador registered a single case on February 27, when in reality, looking retrospectively, there were already 19 undetected cases. Only two weeks later, there were 11 officially registered cases but at least 119 undetected cases (Figure 5). After 54 days, a total of 9,468 positive COVID-19 cases and 474 officially reported deaths, with a sex distribution (55.4% men) that was similar to that reported in China (58% men) and Italy (59.8% men) ^17-19^ The mortality rate for women (3.35%) is almost half of that for men (6.86%). The median age was 42 in men and 39 in women and the age group most affected was the group from 19-50 years old representing 59.6% of the entire cohort, almost doubling the same age group reported in Italy (24.0%) and very similar to the age distribution from China ^18,19^ The reason behind these trends might be due to the fact that Italy has one of the oldest populations in the world. According to the latest data, Italy has 14 million residents over the age of 65 (22%) with an average age of 45.7 years, while in Ecuador the average population is 26.6 years old ^20-22^

In terms of age, the patients between 0 and 50 years old reported a CFR of 1.6%, compared to 0.4% in Italy, 0.4% in China and 0.6% in Spain from the same age groups ^22-24^ When adjusting for sex, age and the presence of comorbidities, mortality increased significantly among elderly men, which is consistent with other regional studies ^18,25^.

The existence of comorbidities is linked with augmented age and therefore increased risk of mortality in COVID-19 patients. The age of patients reporting comorbidities in Ecuador was higher than those without comorbidities. In terms of risk, patients with comorbidities had a CFR% of 10.3% in women and 16.9 % in men, higher than those without comorbidities and in both sexes CGF% averaged 4%. These findings are equivalent to previous studies that have shown that the presence of comorbidities increases the risk in COVID-19 patients to be admitted to the ICU or die due to this disease ^25,26^.

In terms of ethnicity, it is important to point out that self-identified Montubios and Indigenous had a CFR% of 14% and 9% respectively, which is surprisingly higher than Mestizos (6%) and other ethnic groups living in Ecuador (Figure S9-Appendix 1). This is probably due to reduced healthcare access for vulnerable groups ^27^. For influenza, ethnic minorities have the highest estimated fatality rate, most likely due to their social determinants of health, social inequalities and reduced access to health care, especially in rural areas ^28,29^

Although the clinical features of COVID-19 are widely studied in moderate and severe hospitalized patients, information on patients with a less severe symptoms is scarce. We collected self-reported data on symptoms from a sample of patients in home isolation. 53% presented with fatigue, 43% headaches and 42 % a dry cough which is in agreement with studies from other settings. However a higher proportion of patients reported ageusia (37%) and anosmia (36%), when compared to studies from China (5.1% and 5.6%, respectively) and Italy reporting anosmia in 19.4% of patients ^2,30^.

One of the main limitations in understanding the development of COVID-19 in Ecuador is the lack of molecular testing capabilities ^31^. In January, alarms went off with the first suspected COVID-19 case in Ecuador, alarms that denoted the poor readiness of the public health system that took more than 15 days to rule-out a highly suspicious COVID-19 patient, who died with the diagnosis of hepatitis B and atypical pneumonia ^32,33^. At the beginning of the outbreak, the World Health Organization (WHO) emphasized the importance of testing capabilities in order to improve case tracing and diseases detection worldwide (Figure S8-Appendix 1). Nevertheless, Ecuador has not enough capabilities to perform molecular diagnosis (RT-PCR), limiting epidemiological surveillance strategies and case tracing ^15,34^ For this reason, the number of samples taken exceeds the local molecular diagnosis capabilities, causing that thousands of tests are not been reported in a daily basis ^11,12^ Despite the limitations, in some areas of the country, especially richer areas, such as Samborondon, testing rates are as high as those seen in Iceland or even close to those seen in the US (Supplementary Table 3 – Appendix 1) ^35^. This trend might be driven by those patients who were able to pay for their diagnosis and their medical treatment. Therefore, their high attack rate is interpreted as better access to health resources and that is why mortality is also low among these cantons. Although social status and monthly income was not assessed, in Ecuador, blue collar workers have less monetary income than health care and white-collar workers ^36,37^. In Ecuador, it is hypothesized that labor workers and the unemployed have less access to health services, therefore the high case fatality rates among these groups is significantly higher than those in more privileged positions such as politicians, medical doctors or health workers (Supplementary Table 2 – Appendix 1). These results could indicate that poor working conditions, poverty and therefore limited access to health services could be linked to the high mortality rates reported among the most vulnerable groups, a situation described previously ^38^.

Low testing capabilities and high number of suspicious COVID-19 deaths likely distort the calculation age-specific attack and mortality rates across cantons. We found a median delay of two days between the day of sampling to the day of notification and the low number of reported tests (test per million people and the overall count) in the region ^15^. This impacts the ability of contact tracing and other prevention strategies to interrupt SARS-CoV-2 transmission^39^.

Analyzing the years of life lost due to SARS-CoV-2 is challenging due to the uncertainty around the length of the different phases of the disease and the clinical spectrum of the severity of the disease. These uncertainties make it difficult to estimate disability weights that are a crucial component for the DALY calculation. In our study 99.5% of the burden was attributable to the years of life lost due to premature mortality among the study population, with an average of 25.4 YLL per death, and 1.27 DALY per each case of COVID-19. Even though COVID-19 related deaths are higher among elderly populations in developing countries with weakened health systems, mortality is also overstretched among younger populations^40^.

The main limitation in this analysis is the lack of specific data regarding the type of comorbidities and the presence of clinical manifestations during the initial evaluation. This limitation is also evident when reviewing the follow-up file, which includes only the date of death and the date when the case was discharged and closed (Appendix 1).

In conclusion, this is the first epidemiological study of the socio-demographic distribution of COVID-19 in Ecuador and one of the very few reported in Latin America. The results demonstrate the vulnerability of the health system to contain, mitigate, treat and adequately diagnose this type of new viral disease that spread across the country at a speed that exceeds the speed of response.

## Data Availability

Data is included as part of the submission

